# Exploring the emerging concept of precision rehabilitation: a qualitative study

**DOI:** 10.1101/2025.07.03.25330802

**Authors:** Annie Pouliot-Laforte, Evemie Dubé, Dahlia Kairy, Danielle E. Levac

**Affiliations:** Department of Physical Activity Sciences, Faculty of Sciences, Université du Québec à Montréal (QC), Canada; Technopole in Pediatric Rehabilitation, Azrieli Research Center UHC Sainte- Justine, Montreal (QC), Canada; School of Rehabilitation, Faculty of Medicine, University of Montreal, Montreal (QC) Canada; Center for Interdisciplinary Research in Rehabilitation of Greater Montreal – Site IURDPM, Montreal (QC), Canada

**Author notes:** Corresponding Author: Annie Pouliot-Laforte, PhD Technopole in pediatric rehabilitation, Azrieli Research Center UHC Sainte-Justine, Montreal (Qc) Canada; Department of Physical Activity Sciences, Faculty of Sciences, Université du Québec à Montréal (Qc) Canada.

**Keywords:** Rehabilitation, Personalization, Precision rehabilitation, Precision medicine, Qualitative Study, Interviews, Consultation, Scoping study

## Abstract

**Purpose:** This descriptive qualitative study explored stakeholders’ perspectives on precision rehabilitation concepts, barriers, facilitators, and future directions as part of a convergent mixed methods scoping review.

**Materials and methods:** Sixteen clinicians, administrators, and researchers from three North American tertiary care rehabilitation centers were recruited using convenience and snowball sampling to participate in individual semi-structured interviews. Conventional qualitative content analysis followed a deductive thematic approach based on predetermined categories.

**Results:** Analyses revealed three main themes: 1) Although precision rehabilitation shares foundational concepts with precision medicine, there are certain elements, such as personalization, that are uniquely expressed; 2) Rehabilitation-specific facilitators to precision approaches include the use of unobtrusive technology to collect large amounts of data in real-world contexts, while barriers include rehabilitation’s typically small, heterogeneous sample sizes; and 3) The future of precision rehabilitation will require collaborative data-sharing to focus on determining care trajectories that enhance functional outcomes.

**Conclusion:** Findings provide the first qualitative synthesis of stakeholder perspectives to complement quantitative evidence and inform the emerging field of precision rehabilitation.

## 1. Introduction

Precision medicine integrates individual biological, environmental, and behavioral data to guide diagnosis and treatment (Schleidgen et al., 2013). Precision approaches have gained substantial momentum across many medical specialties, enabling early diagnosis and tailored interventions (Adams & Petersen, 2016). Progress in precision medicine is fueled by health data digitalization, advances in genomic sequencing, wearables that enable continuous real world data collection (Gambhir et al., 2018), and artificial intelligence (AI) models trained on large databases that can identify treatment responders and non-responders, recommend optimal interventions, and adaptively modify treatment parameters in real time (Johnson et al., 2021). While implementation barriers exist, such as data standardization challenges, precision medicine is an established paradigm with substantial research funding and policy support across the healthcare ecosystem.

In contrast, precision approaches in rehabilitation remain significantly less developed. Rehabilitation’s focus on promoting function across the lifespan through addressing impairments, activity limitations, and participation restrictions (World Health Organization, 2023) presents unique opportunities and challenges for precision approaches. These intersecting domains require consideration of numerous variables that influence rehabilitation outcomes, making the development of precision frameworks particularly challenging (Lin & Stein, 2023).

Current conceptualizations of precision rehabilitation are grounded in precision medicine principles yet acknowledge the unique aspects of precision as applied to rehabilitation practice. In 2022, French et al. identified the core objective of precision rehabilitation as consistent with its medical counterpart: delivering the right intervention, at the right time, to the right individual (French et al., 2022). However, they described precision rehabilitation as focusing on preventing functional decline and maintaining functional independence, emphasizing that precision rehabilitation centers specifically on function at an individual level, encompassing not only physical aspects but also cognitive and psychosocial dimensions (French et al., 2022). The editor of a 2022 special issue of the Journal of Physical Therapy devoted to precision rehabilitation provides further conceptual clarification by describing precision rehabilitation as "linking behavior to biology to deliver patient-centered care" (Jette, 2022). The editorial positions physical therapy at "the intersecting frontiers of biologic, behavioral and population health research," creating an ideal environment for precision rehabilitation to flourish. This perspective emphasizes the integrative nature of rehabilitation as a discipline uniquely positioned to advance precision approaches.

Cotton et al. (2022) identified critical needs to advance precision rehabilitation, advocating for a unifying transdisciplinary framework to guide both research and practice (Cotton et al., 2022). They argued that such a framework should prioritize functional, long-term outcomes identified by patients themselves. Additionally, they emphasize the importance of exploring how context variations across heterogeneous patient groups affect treatment efficacy. Their recommendations include enhancing the diversity of patients in research studies and incorporating genetic and genomic factors into rehabilitation approaches. Cotton et al.’s recent work (2024) proposes a comprehensive causal framework for precision rehabilitation that takes into account the complexity of rehabilitation science (Cotton et al., 2024). The framework aims to identify Optimal Dynamic Treatment Regimens (ODTR), which are decision-making strategies that use available measurements and biomarkers to identify interventions likely to maximize long- term function, providing an example of how causal models can learn from heterogeneous data from different sources.

To supplement these important efforts in advancing precision rehabilitation, it is important to understand how rehabilitation stakeholders conceptualize precision concepts in real-world clinical settings. Exploring the perspectives of those who will ultimately implement and benefit from precision rehabilitation will help to bridge the gap between research and practice to inform implementation initiatives.

### Study aims

This study is the qualitative component of a convergent mixed methods scoping study (Pouliot-Laforte et al., 2025) aiming to describe and map the current state of knowledge regarding precision rehabilitation. The objective is to explore how stakeholders in the field describe precision rehabilitation, its current and future state, and its barriers and facilitators.

## 2. Methods Design

Qualitative description aims to provide a comprehensive, straightforward description of an experience or phenomenon in everyday terms (Doyle et al., 2020; Sandelowski, 2010). We anchored this qualitative component in the consultation phase of the scoping study process (Arksey & O’Malley, 2005; Levac et al., 2010), and followed Buus et al, 2022’s recommendations to maximize scientific rigor (Buus et al., 2022). We report our findings according to the Consolidated Criteria for Reporting Qualitative Research (COREQ) guidelines (Tong et al., 2007). The study was approved by the Research Ethics Board of CHU Sainte-Justine Research Centre (no. 2024-6324). All study participants provided informed consent prior to their participation.

### Recruitment

We recruited participants from three large North American tertiary care rehabilitation centers using convenience and snowball sampling. A designated contact at each site identified potential researchers, clinicians, and managers, then emailed them study information. Interested individuals contacted our team directly to participate. Prior knowledge of or experience with precision rehabilitation was not required.

### Data collection

Participants completed a standardized demographic form (including profession, gender, age, country and region, place of employment and years of experience) prior to participating in a 30–45-minute semi-structured individual interview in-person or via a secured virtual platform (Zoom, Zoom Video Communications, San José, USA). Interviews were conducted by two members of the research team with qualitative research expertise (A.P-L. and D.L.) using an interview guide centered on four questions:

1) What does precision rehabilitation mean to you? 2) How would you describe precision rehabilitation in your context? 3) What are the difficulties/obstacles that you have encountered or anticipated to encounter? 4) Based on your experiences, what is the role of artificial intelligence and interactives technologies in precision approach? Interviewers followed each question with targeted probes based on participants’ responses.

### Data analysis

Interviews were audio-recorded, de-identified, transcribed verbatim, and analyzed using a two-step hybrid deductive-inductive thematic approach (Patton, 2014; Vaismoradi et al., 2013) to identify patterns within data (Braun & Clarke, 2006). Following deductive guidelines (Fife & Gossner, 2024), coding was structured around four predetermined themes from the interview questions: 1) Current Situation: problems addressable by precision rehabilitation, 2) Key Concepts: precision rehabilitation concepts and their relationship to precision medicine, 3) Barriers and Facilitators: challenges and enablers to precision rehabilitation, and 4) Future Directions: next steps and vision for rehabilitation under precision principles. While initially analyzed separately, facilitators and barriers were later combined when we observed that concepts positioned positively functioned as facilitators, while their absence or negative positioning constituted barriers. Sentences served as the coding units with text segments were labeled with corresponding codes. Subsequently, one transcript was independently coded by A.P-L. and D.L. The coded transcript was then revised collaboratively to reach an inter-judge agreement. All transcripts were then coded by A.P-L and D.L.; the coding team met regularly to discuss discrepancies, reach consensus, and refine understanding of the codebook as needed. Upon completion of the coding phase, A.P-L revised all the units of analysis of each code and synthesized the material. Following the coding phase, we used an inductive approach to identify sub-themes. Illustrative citations of the themes and sub-themes were identified in the transcripts. Three participants also engaged in reviewing portions of their data to verify accuracy.

## 3. Results

The sociodemographic characteristics of the 16 interview participants are presented in Table 1. From the qualitative analysis, three themes emerged, two of which had 3 sub- themes: 1) Key concepts of precision rehabilitation parallel those of precision medicine but have their own unique features, 2) There are rehabilitation-specific facilitators and barriers to a precision approach, 3) The future of precision rehabilitation will focus on function. Figures 1 and 2 illustrate the themes and subthemes.

**Figure 1.**
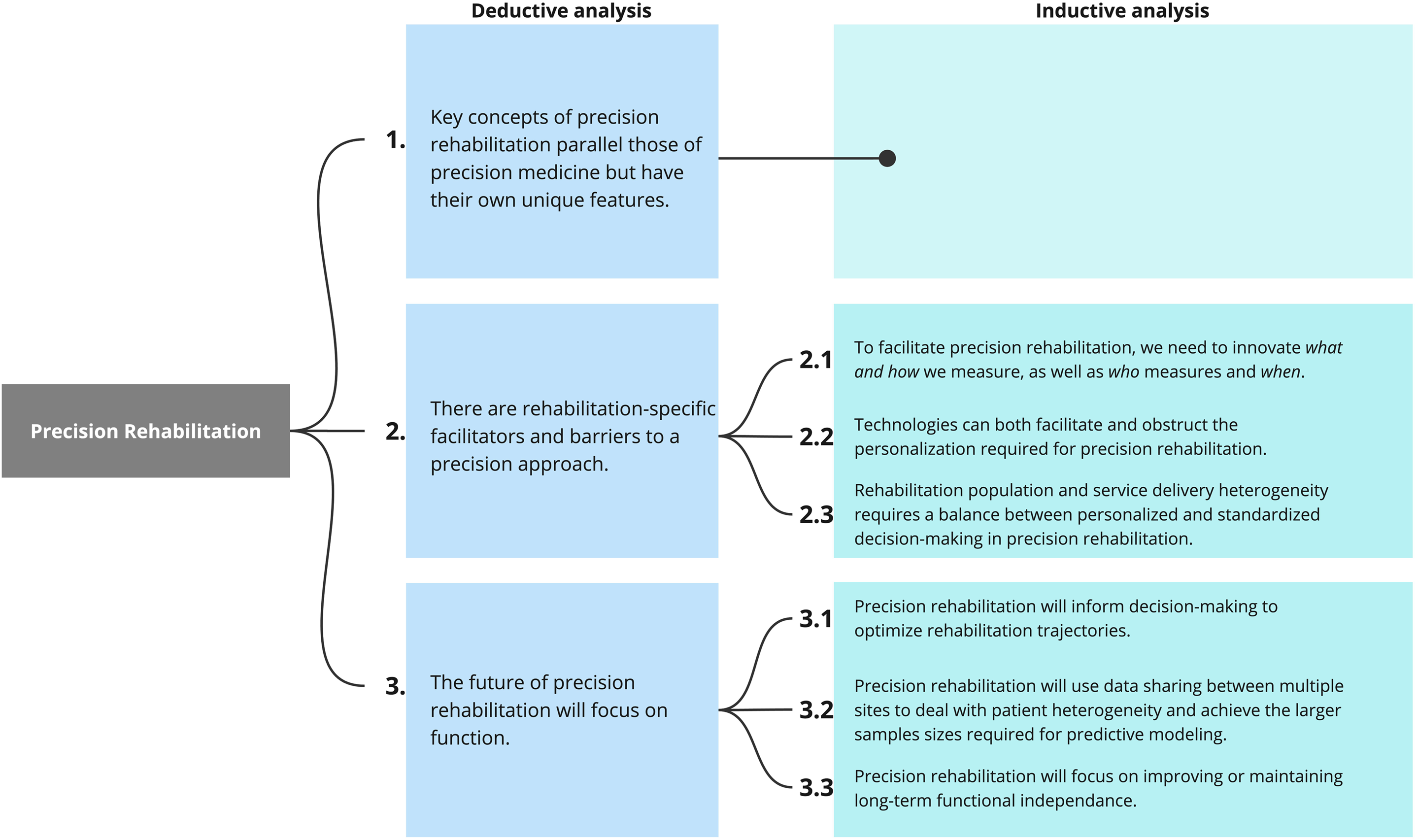
Deductive and inductive thematic analyses The deductive analysis identified three main themes characterizing precision rehabilitation (blue rectangle): (1) key concepts aligned with precision medicine, (2) specific facilitators and barriers inherent to rehabilitation practice, and (3) a functional focus for the future of precision rehabilitation. Inductive analysis expanded these themes into detailed subthemes represented by green rectangle.

**Figure 2.**
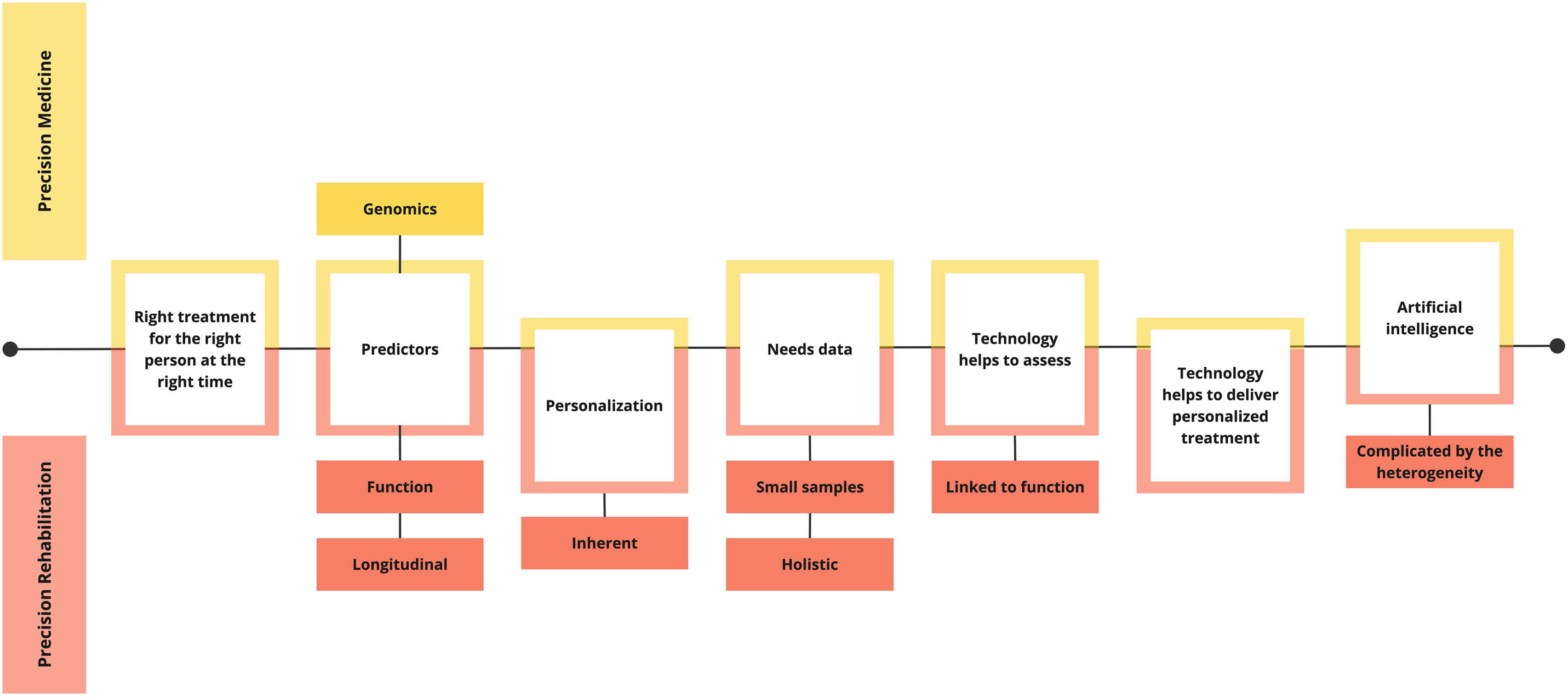
Key concepts conceptual comparison of precision medicine and precision rehabilitation. The conceptual map illustrates similarities and distinctive characteristics between precision medicine and precision rehabilitation. Shared foundational principles are presented along the mid-line. Precision medicine specific aspects (yellow boxes), including genomics, contrast with precision rehabilitation specific elements (orange boxes) emphasizing function, longitudinal assessment, holistic perspectives, small sample considerations, and challenges introduced by heterogeneity.

**Table 1.**
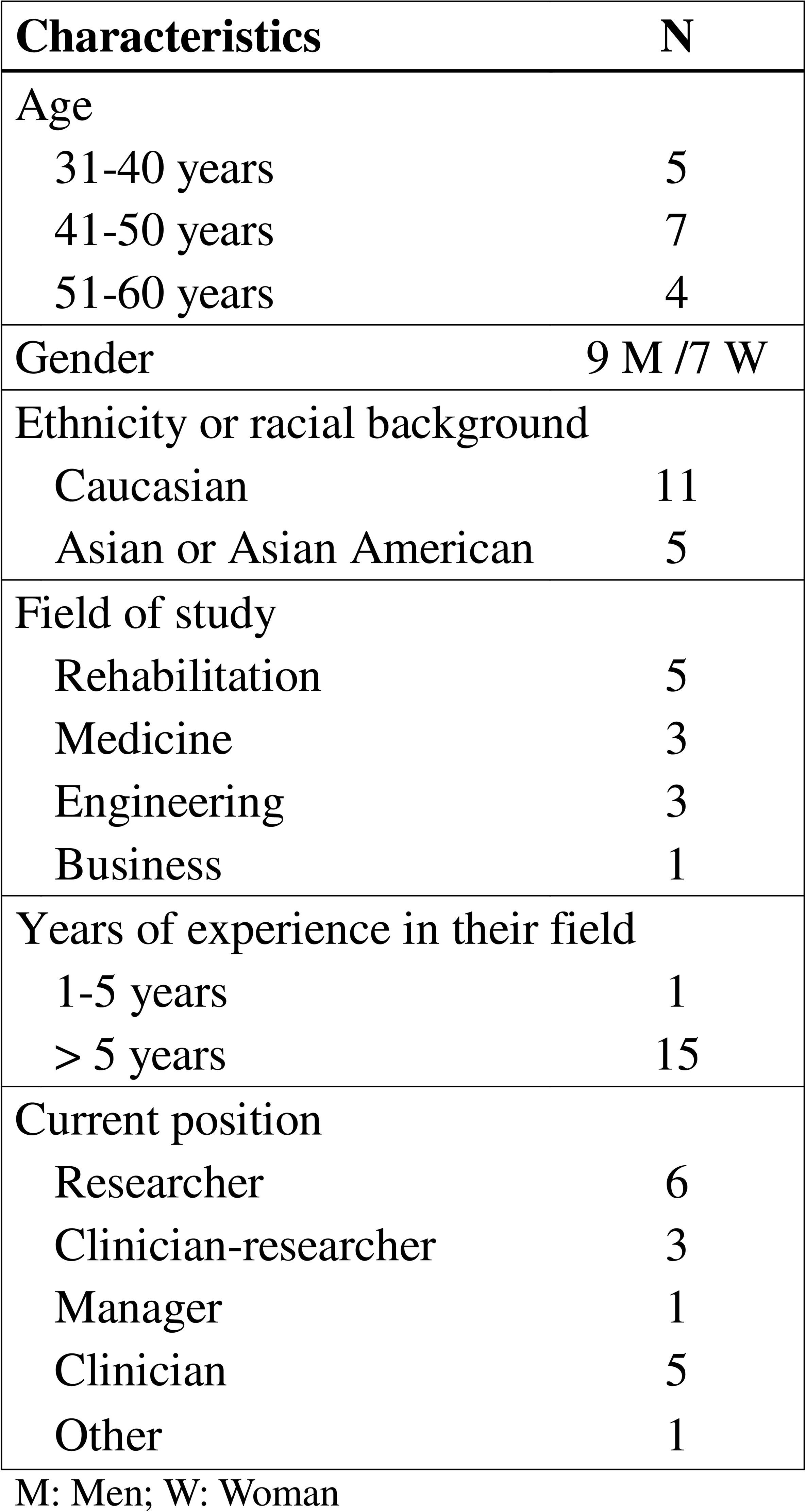
Participants’ sociodemographic characteristics.

**Theme 1: Key concepts of precision rehabilitation parallel precision medicine but have their own unique features.** Participants described their understanding of how precision rehabilitation shares key concepts with precision medicine, including its general definition and the importance of biomarkers and personalization in a precision approach. Firstly, participants defined both precision medicine and precision rehabilitation using words identical to, or closely resembling, ‘*the right treatment at the right time for the right person*’. Participants also described predicting and categorizing as key elements of precision rehabilitation. For example, one participant said: *“[Precision rehabilitation is] …to be able to come up with a much better understanding of what worked for what child, based on what characteristics, [to inform] future treatment.”* (Participant 4).

Another stated: “[…] *ideally before they get into an intervention, if we can categorize who would benefit from what, is in my mind, precision.”* (Participant 1).

Secondly, participants identified biomarkers as a central concept in any precision approach. Participants described the need for rehabilitation-specific biomarkers to identify intervention responders and non-responders. In this regard, several participants viewed the genomic biomarkers typical of precision medicine as equally relevant to rehabilitation. For example, one participant said: “*I do see precision health (using genetics) informing my understanding of the child at an individual level and how it may impact and guide rehabilitation strategies. So let me just give you a really high-level example. So, within all of our [name of diagnosis] cohorts, we have children that when we do genomic testing, that instead of having [diagnosis], they have [diagnosis]. So, there’s a precision health test that I can do […] that tells me as a clinician that I really need to make sure I’m not doing anything to weaken that child because that child is going to have a different picture to the typical child with [diagnosis] - that they’re going to have more weakness, and they may have a progression. So, there’s probably a lot of elements of precision health and how that’s going to inform rehabilitation strategies that we need to really capitalize on.”* (Participant 10). However, others noted that more emphasis should be placed on non-genomic biomarkers based on function. For example, one participant said: *“And so obviously in rehabilitation, we wouldn’t be as focused on genetic markers and other kinds of biomarkers, but we might be more focused on using clinical outcome measures.”* (Participant 13). Another said: “*I know, in the literature, there’s much more of an epigenetic focus. We don’t really consider that too much because*

*we know that children are changing and developing and then that pathway is unique for each child as well.”* (Participant 7). These biomarkers of functional status could be used in prediction models: *“So that would be a beautiful thing to just have a marker of function over time and similar ones for communication, for manual ability, for eating. But I also think it should go broader, so I would love to have a marker for well-being, for participation, and then I also think pain is a really important marker in terms of negatively impacting on quality of life and well-being, so having a marker for pain.”* (Participant 10).

Thirdly, participants identified the concept of ‘personalization’ as central to both precision medicine and rehabilitation, but as requiring unique interpretation when applied to rehabilitation. Specifically, participants viewed personalization as an essential, inherent component of any rehabilitation service delivery, unrelated to a precision approach. For example, one participant said: “*And I mean it probably it’s something that’s closely tied to rehabilitation in many instances anyway, because I mean that the care is really personalized to the individual in terms of their goals and their impairments.”* (Participant 8). Another mentioned: “*Yeah, I mean I think we’ve been kind of doing that all along. Because we recognize you know that the children are so unique, each and every one right now in our work. I mean, I can’t think of two kids who have been, you know, alike in that sense. […] I think we’ve had that kind of mindset all along because we’ve never, you know, deluded ourselves to think that, oh, yeah, this is a solution that’s going to work for, you know, hundreds of kids, right?”* (Participant 7).

Participants suggested ways to reconcile the concept of personalization in a precision rehabilitation approach, including that it could provide a structure to systematize what is currently an unstandardized, clinician-specific personalization process. For example, one participant said: “*I think maybe it’s systemizing it a little bit more because I think that individual clinicians are always personalizing to the child. But I mean each individual clinician will have sort of their knowledge base. But if we want to be able to provide a standard of care across clinicians and organizations then it has to be like systemized in a way that we can provide evidence-based recommendations to how you tailor and personalize.”* (Participant 9). Participants also suggested that a precision approach to personalization could help include data-informed evidence alongside considerations of client factors in rehabilitation decision-making. One participant reflected: “*I think client centered goals are obviously important and that’s something that we concentrate on quite a bit. When I think about precision, I guess whereas client centered is based on client goals, the precision may be more based on research and kind of best practice which don’t always align with client-centered goals.”* (Participant 11). Participants agreed that artificial intelligence (AI) models are as essential for personalization in precision rehabilitation as they are in medicine: *“So we’re absolutely going to need AI in this. If we’re going to have the large data sets, we’ve got to have some way of being able to pull things out, to be able to look at patterns is that’s the only way, I mean. We’re not going to be able to do it otherwise. And I mean I think that’s things that we do as clinicians when we look at kids, I mean. That’s what we’re doing all the time is trying to pull out those patterns, but of course we can’t do it effectively or particularly accurately.”* (Participant 4).

Lastly, participants reflected that the word ‘precision’ itself may be less well known in rehabilitation as compared to medicine. They noted that rehabilitation tends to use other words to describe the need to provide optimal and timely interventions, such as “care efficiency.” For example, one participant mentioned: *“No one ever uses the word precision rehabilitation and … it’s more like hey we have this amount of time what can we get done. It’s more about, I would say like moving people through the healthcare continuum is generally the talk versus precision rehabilitation.”* (Participant 15). Another said: “*Not the term necessarily, but that like. in […mention of a healthcare center…], there’s a big push for like, you know, the right service, right time, right patients. Those are kind of concepts that are ingrained. But I’ve never heard it referenced as precision.”* (Participant 11).

Participants provided several examples when asked to describe elements of their current research or clinical practice that aligned with their understanding of precision rehabilitation, including using technologies to improve assessment timing, genomic testing to inform decision-making, and psychosocial factors to predict outcomes. Table 2 categorizes these examples according to the concepts of precision rehabilitation that emerged in Theme 1.

**Table 2.**
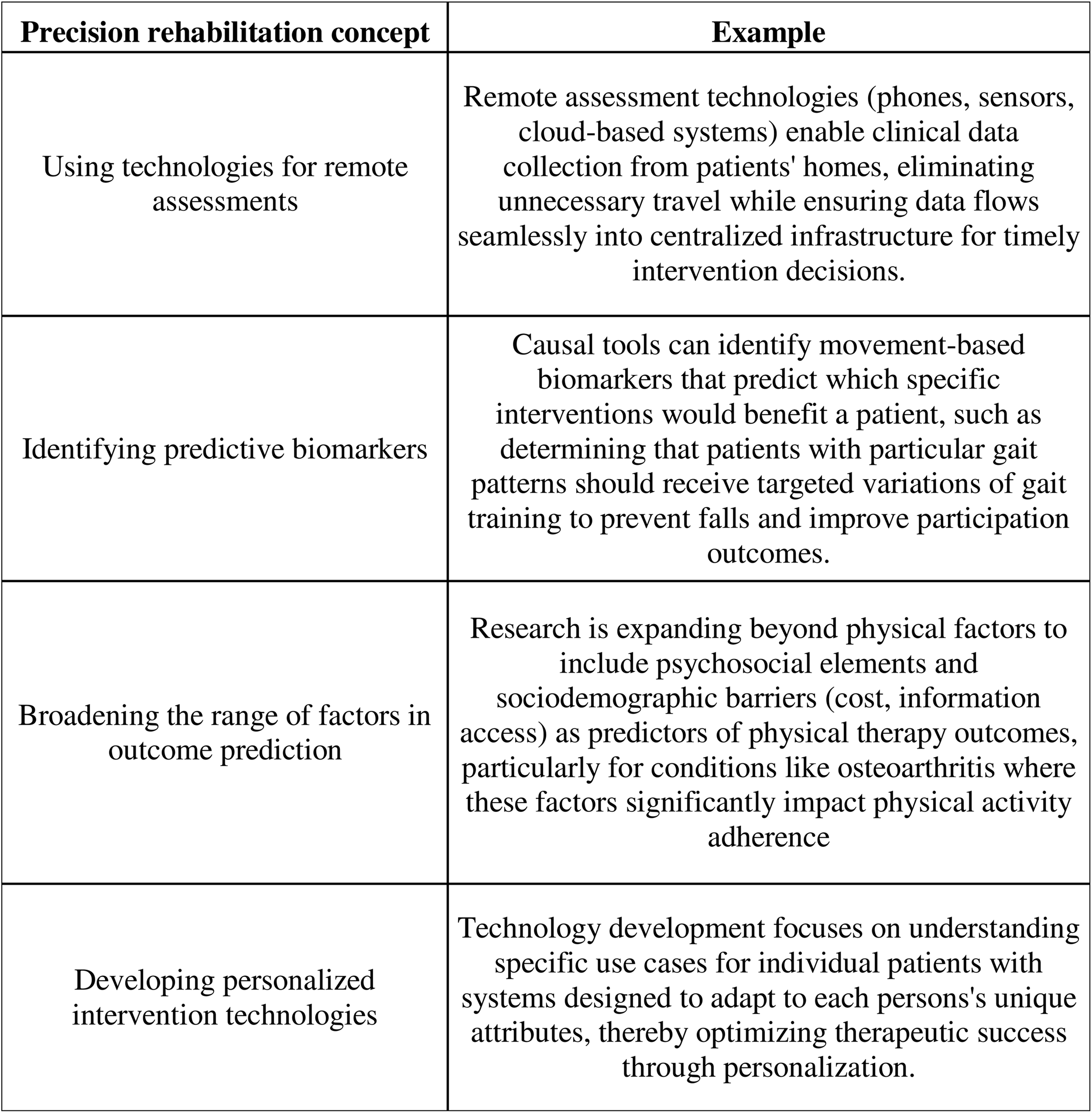
Examples of current initiatives aligning with a precision rehabilitation approach.

**Theme 2: There are rehabilitation-specific facilitators and barriers to a precision approach.** Three subthemes summarize participants’ perspectives on facilitators and barriers to precision rehabilitation. Within each subtheme, the same concept emerged as a facilitator (when present) or as a barrier (when absent): 2.1) To facilitate precision rehabilitation, we need to innovate *what and how* we measure, as well as *who* measures and *when*, 2.2) Technologies can both facilitate and obstruct the personalization required for precision rehabilitation, and 2.3) The heterogeneity of rehabilitation populations and interventions complicates decisions based primarily on evidence.

### 2.1 To facilitate precision rehabilitation, we need to innovate *what and how* we measure, as well as *who* measures and *when*

Participants agreed that more frequent measurement and data collection will facilitate precision rehabilitation. With respect to **what** we measure, participants agreed that the current lack of measurement of psychosocial factors influencing decision-making and outcomes is a barrier to precision rehabilitation because it means that these factors will not be considered in decision- making models. For example, one participant said: *“I don’t think we’ve been as holistic as we might need to be to capture everything relating to physical and psychosocial. And you know, cognitive characteristics of the child, like the whole child. So, I think we’re, you know, we’ve got a lot of work to do to find that best set of things to measure and figure out when to measure them.”* (Participant 4). Another participant stated: “*[…] the psychosocial part plays a big role in someone’s response to treatment. And so, when I started out, that was one of the biggest challenges. My PhD was in a research lab where everything’s clean, it’s just you’re working on cells. But when you start looking at the human, it’s very different. And patients have variability, patients have perceptions about treatments, and so it’s just not as easy to just purely look at biology, in that respect.”* (Participant 3). To support psychosocial factor measurement, participants identified research facilitators such as greater use of qualitative research methodologies. For example, a researcher noted: “*I’m leaning more towards those single case experimental designs. That where, you know kids are serving as their own controls and we can sort of try to understand those individual experiences in a little bit more depth. Through repeated measures and more continuous monitoring, as well as through mixed methods and really trying to merge that the quantitative data with the qualitative data because I don’t think you can’t capture it all through quantitative data and you know quantitative data has its own subjectivity to it.*” (Participant 9). Yet participants recognized the challenge in identifying which psychosocial factors to measure to avoid increasing sources of information and complicating decisions. For example, one participant said : “*I think forward progress in the field will be very much helped by people who perhaps have a simple vision on what might be the important things to try to assess, because there’s going to be a mass of data and if people aren’t coming in with that pragmatic approach on how really focus on things that are meaningful to the families, I think we’re going to get lost. I think we’ll have these masses of data, and we won’t really be able to get things that are useful from them.”* (Participant 10).

With respect to **when** we measure, participants identified that current assessment timing is a barrier to identifying optimal intervention moments. Measurements currently happen at rigidly pre-determined intervals (for example, yearly clinic visits) that often have little relevance to clients’ needs. For example, a participant stated: *“It is hard - sometimes like impossible - for families to get here. Like there’s nothing worse than bringing a family in for a clinic visit and they’ve traveled 8 hours, they’ve stayed overnight, and they’ve arranged babysitting or whatever for their other kids. And they’re missing school. And it’s just like, yeah, everything is fine. There’s nothing for us to do, like, really, if you’re bringing them in person, you want to capitalize on that.”* (Participant 14). This timing can impede opportunities to intervene at more appropriate times. For example, one participant said: “*I think for a few of the conditions that I work with, we don’t really actually know what the windows are. I’m thinking like with our kids with cerebral palsy, we get referrals for them for knee flexion contractures, for plantar flexion contractures and you know, they’re 5-6 years old. And then the question is always like, is the surgery going to need to be repeated? What is their growth going to be? I mean trying to have more longitudinal studies that look at exactly these kinds of things.”* (Participant 14).

With respect to **who** measures, participants identified relying on clinicians to assess and collect data is a substantial barrier. For example, one participant said: “*Collecting the data consistently is the problem, right? Just be clear the data is hugely problematic in terms of clinicians. You’re going to be relying on clinicians to collect data. They’re already time stressed. They don’t necessarily have the tools that they need to, they may not have the same goals.”* (Participant 9). Participants identified two facilitators to support clinicians in data collection. The first is greater institutional support to emphasize the importance of data collection and making it a priority at an institutional level. For example, one participant said: *“You know when it’s some way strategic priority, then there’s then it’s more likely to get done. Clinicians will be more likely to be given time and rewarded for participating in this sort of thing, and I think it also carries down to even how you know positions are filled and things like that because you know if it’s ingrained in the culture of an organization that this is what we do when they’re interviewing for clinical placements and things like that.”* (Participant 9). The second facilitator relates to ***how*** we measure. Participants identified that technologies could facilitate unobtrusive collection of large amounts of data in real world settings, reducing the burden on clinicians for data collection. For example, one participant said: “*But time is such a barrier, right? I mean, whenever I suggest to clinicians, why don’t we do this? It’s always the time barrier, can’t do it, can’t fit it in. I think if there are things that can be fitted in again within the context of the family, the child, the parent, you know, wearables again. Smartphones, like tracking things that we can do, I think. […]*” (Participant 4). Another said: “*I think that’s where technology will help us because we won’t have to do everything all at once anymore, either right? Like certain things you could capture as they go along through their day, which is more meaningful anyway. You could get their six-minute walk test some other way. You don’t have to do it here at the centre… maybe don’t even have to do the GMFM with the kid after he’s done 3 hours of gait lab. We may have gotten comprehensive assessments, but the child certainly wasn’t at their best, they weren’t representative of what they could do.”* (Participant 4). A participant stated: “*My vision, for example, to instrument a 10-meter walk test is actually just to make their assessments easier, right? Right now they have to go get a stopwatch. Like if someone needs assistance, they might need someone else, you know there by their side. Whereas if we have a such an area where you basically swipe someone’s wristband, just have them walk and then you got a 10-meter walk test and then you happen to also get all this gait data. Then, you actually ask less of the clinician. So, and then the same with the smartphone system designed to be super easy to use. And so, I think that’s really important for these new technologies, not actually creating burdens.”* (Participant 3).

### 2.2 Technologies can both facilitate and obstruct the personalization required for precision rehabilitation

Using technologies for data collection can facilitate personalized decision-making by providing us with more detailed information that would otherwise be difficult to measure or obtain. For example, one participant noted: “*Continuous data capture, for example from a fabric that kids can wear all the time, can help us make better decisions.”* (Participant 7). Another said: *“We track their smartphone to see how much they walk, how many steps they get? Are they in the community? We do heat mapping which means are they going to the movies? Or are they eating out at restaurants?”* (Participant 1).

However, when using technologies as intervention modalities, participants identified the decision-making burden of technologies that offer multiple parameters as a barrier. Being able to customize technological tools, such as games and apps, often requires training and setup and calibration time, which was identified as a barrier because it can detract from valuable therapy time. For example, one participant said: *“Sometimes personalizing games like that and calibrating for that can take some time and that can be a bit of a turn off. (The time it takes to reach the treatment area where the technology is located is) going to cut our time down to 30 minutes and then it’s going to take maybe 5 minutes for me to customize”* (Participant 12). Additionally, the lack of resources and guidance on where to start when personalizing technology for individual patients was identified as a barrier making technology use more daunting and less efficient. For example, one said: *“Sometimes it’s kind of just like, OK, let’s start here and you are kind of like stumbling in the beginning, because you are problem solving […]. But where is the right starting point for that person? We might start too hard or too, you know, too easy. So, some additional guidance or support for personalizing would be great”* (Participant 11).

Rehabilitation interventions need to be personalized, but at the same time applicable to more than one person. Participants identified that attaining this balance between personalization and generalization is a barrier to technology use in a precision rehabilitation approach. Participants described how they appreciate tools that can be tailored to meet one person’s specific needs, but that to justify the learning and time required in their use, they also need the tool to be generalizable enough to work for many people. For example, one participant said: “*There are different, very different, goals that these kids are trying to achieve, and clinicians are working towards. So having a tool that kind of meets all those needs is a challenge. And if you want this to be a useful tool, you just ideally want one system that could do it for all, but at the same time have the ability to individualize itself to the goals of the particular person, right? So that’s part of the challenge I think.*” (Participant 8). The issue of balancing personalization and generalization is also a barrier for bringing personalized rehabilitation technologies to market. A participant with experience in this area described this challenge as follows: “*You need something that’s going to address the needs of a larger group. It can still be individualized, perhaps, but it needs to be individualizable to a large enough group so that it is commercially viable.*” (Participant 8) The participant went on to speak about how this can complexify the design and development of new technologies: “*So we’ll design a technology and we’re trying to keep things simple so that you know we can do it with finite time and resources. But then we start thinking about, I guess, like every child that we try it with, and we learn something from, we start adding features to the technology. So, feature creep I guess and then things can start to get more complicated*.” (Participant 9).

### 2.3. The heterogeneity of rehabilitation population and interventions complicates decisions based primarily on evidence

Participants described how the inherent variability in both rehabilitation populations (for example, patients with stroke) and interventions (for example, gait training) is a barrier to precision rehabilitation, because data-driven predictive algorithms that inform decisions about individual care might have been built on a population that doesn’t necessarily resemble the individual client. For example, one participant said: “*So, we’re assuming that an intervention is an intervention, but the intervention is so dependent, as we all know, on the provider and how they do it. I mean, we’re not providing even the same intervention. So, for us to think that our interventions are as simplistic as you know, this will help us know whether we should be doing X or Y. It’s not like drugs, right? It’s so much more complicated than that. We don’t know the characteristics of those interventions that are going into that mix of helping us decide what’s working, what isn’t, and how that fits with the individual.”* (Participant 4). Another said: *“We don’t really have a standard. I think it’s very specific to the clinician’s expertise around how like their clinical reasoning of, based on the assessment that they take, yeah.”* (Participant 12). Participants noted that this lack of clarity in intervention content exists at the level of rehabilitation evidence: “I *think one thing that’s often overlooked is really detailed in that documentation of the interventions, right? Like every time Cochrane reviews tries to do a meta-analysis say like, well, this was futile because no one said what they did.”* (Participant 2). To facilitate precision rehabilitation, participants noted the need to systematize how decisions are made and to study the process of systematization. For example, one participant said: “*If everybody is getting a different intervention, how do you measure that sort of thing? So that’s where it comes back to systemizing - there has to be a reason that you chose this set of tools to apply to this child. You’re researching that systematic way of choosing the menu of options… you’re not necessarily like measuring a specific intervention, so much as the way that you’re prescribing the intervention and is that way that you’re prescribing the intervention leading to good outcomes.*” (Participant 9).

Indeed, participants described the focus on using information from big datasets to make decisions about individual patients as a barrier to implementing precision rehabilitation. They struggled with how to balance using evidence versus individual client factors to inform decision-making. For example, one participant said: “*Whereas client centered is based on client goals like the precision may be more based on research and kind of best practice which don’t always align with client-centered goals. So that’s kind of yeah, it can become tricky in that way.”* (Participant 11). Participants agreed that the goal of using these large databases to make classifications and predictions is important but described concern about doing this in the typically heterogeneous rehabilitation populations. For example, one participant said: *“You know, one of the challenges is like in the area that I work in, we often struggle with getting adequate sample sizes and so often in the research you’ll see kind of very heterogeneous samples.”* (Participant 13). Another said: *“It’s just so hard when everybody is so incredibly different and, like, they have different functional goals that they want to work on and like, I just don’t understand like necessarily how you would like predict that their outcomes and what it would look like in the future.”* (Participant 15). Participants also viewed the ethical challenges of using big datasets to inform decision-making as a barrier to precision rehabilitation. For example, one participant said: “*One of the issues that I see with sort of this personalized medicine approach is that you can… If you come up with a model for a patient immediately when they come into clinic, you can predict how they’re going to do. There’s no potential leeway in their treatment approach, right? If insurance companies, etc, say they’re going to put this patient’s data into a model and we’re going to predict how they’re going to do. Then that removes the ability of the physician to think outside the box. So, if you try and go outside those guidelines, it’s almost certainly not approved. So that does stifle innovation to some extent in patient care and thinking outside the box. And so that’s one of the issues*.” (Participant 3). Another participant said: “[…] *particularly with insurance clamping down more and more…then if you’re just going to put all this patient data in a computer and spits out, this patient doesn’t get this treatment. I’m worried that that will leave certain people behind who can’t afford maybe certain treatments or will be denied by the insurance companies.*” (Participant 3).

**Theme 3. The future of precision rehabilitation will focus on function.** Three subthemes reflected participants’ visions of the future of precision rehabilitation: 3.1)

Precision rehabilitation will inform decisions and optimize rehabilitation trajectories, 3.2) Precision rehabilitation will share data between multiple sites to deal with patient heterogeneity and achieve the larger samples sizes required for predictive modeling, and 3.3) Precision rehabilitation will focus on improving or maintaining long-term functional independence.

### 3.1. Precision rehabilitation will inform decisions and optimize rehabilitation trajectories

Participants viewed the future or precision rehabilitation as providing information for decisions that better support rehabilitation care trajectories. For example, one participant said: *My hope is when we input a person’s information, it will guide you on what they should be doing.”* (Participant 3). Another participant said: “*We need to look at more creative ways to meet the demands of our growing population, and I think that’s where precision rehabilitation has its niche is because you need to be able to be: “OK I don’t need to see this person right now”. You need to somehow eliminate visits and not just have the structured routine visits on a regular basis. You need a model that allows you to eliminate visits because they’re confident that you’re intervening at the exact time and with the correct people.”* (Participant 9). The participant further stated: *“It would be nice to know: hey, this person on the waiting list, based on these criteria, you know, they’re the perfect candidate right now for this dose.”* (Participant 9).

### 3.2 Precision rehabilitation will share data between multiple sites to deal with patient heterogeneity and achieve the larger samples sizes required for predictive modeling

Participants described data sharing as essential to the future of precision rehabilitation. They noted that this will require standardizing data collection across multiple sites. For example, one participant said: *“So, you need volume… to get the volume, then you’ve got to start to have partners who are capturing the same stuff as you at this maybe the same periods of time. I mean you can get away with some of that with modeling, so we don’t need that (to be) identical, but you certainly need to have to measures the same and we know from past history that’s one of the biggest challenges of all is getting that set of best measures.”* (Participant 4). Another participant said: “*I mean access to data I think is a big (problem). So, if we’re going to use things like machine learning, we need access to decent quality data, and I think that’s going to get easier with a lot of the sites like collecting, using kind of more standard data sets.*” (Participant 13). Data volume requires multidisciplinary contributors with different skill sets. For example, one participant said: “T*hese conglomerates, like it’s really essential. We’ve got to have networks of people working together… it’s got to go really broadly across treatment facilities who are pulling all of this together into some sort of big database, right? Where you’ve got the analysts who were able to work with it and being able to build the AI piece of it, right?* (Participant 4). Data sharing is essential to enable build predictive AI models: *“Eventually make it open data, because there’s such a paucity of* (type of) *data. So, we really want to bridge that gap. So yes, to get to a point where, you know, we can take advantage of transfer learning where we have a big pot of data of, you know, kids across you know, many different age ranges and then we can have pre trained models you know instead of having to put so much on every new family that that comes on board. Maybe we can have a pre trained model that is going to be like an average representation of children, you know, like five to seven years old, so they’re not starting from scratch.”* (Participant 5).

### 3.3. Precision rehabilitation will focus on improving or maintaining long-term functional independence

Participants spoke of their vision for the future of precision rehabilitation as improving functional outcomes. To do so, they noted that models must focus on predicting rehabilitation-relevant outcomes, such as participation. For example, one participant said: *“And so that’s exactly why we need sort of computational approaches that go across the levels of the ICF and across time, right? To understand that an intervention we do in inpatient rehabilitation now might actually alter someone’s walking capacity a year from now.”* (Participant 2). And the participant continued: *“All these new technologies for measuring impairments, people are advocating for them, but they really need to be able to link to functional outcomes.”* (Participant 2). Participants viewed a precision rehabilitation future as better accounting for complex relationships between factors, enabling better prediction of functional outcomes. For example, this participant said: *“By capturing a lot more in terms of a diversity of measures, by capturing these electronically, by being able to look at things properly analytically and look at relationships among (variables).”* (Participant 1). Finally, participants noted that an emphasis on using information from technological assessments to predict and optimize participation will be key to the future of precision rehabilitation : “*As people are advancing collecting large data sets and rehab and you know, we’re developing technologies to more precisely monitor qualitatively people’s impairments, we’ll need tools to better understand how our treatment interventions change people at the level of impairment and of course link it across the ICF and change actual participation level outcomes.*” (Participant 2).

## DISCUSSION

Understanding how precision rehabilitation is interpreted in practice is critical to its development. This qualitative study explores stakeholder perceptions of the core concepts, implementation challenges, and enabling factors of precision rehabilitation. Our findings validate current knowledge while revealing new, distinctive aspects that distinguish rehabilitation applications from precision medicine approaches.

While previous work (French et al., 2022; Lin & Stein, 2023) has described the complexity of rehabilitation interventions as a challenge for precision approaches, the results of this study provide specific insights into how stakeholders grapple with this complexity in practice as it relates to personalization. Participants highlighted the challenge of reconciling the data-driven standardization of precision with rehabilitation’s deeply rooted tradition of individualized interventions. Indeed, participants expressed a tension between their desire to prioritize experience-based clinical expertise and the potential opportunities for better decision making offered by the use of large data-driven models. They noted the challenge of incorporating the psychosocial factors that are essential to rehabilitation decisions into standardized decision-making models, as these factors are rarely measured quantitatively. They also identified ethical concerns about how predictive models might limit clinical innovation. These findings are consistent with those reported by Rosenberg et al. (2022), who underscored the necessity for ‘individual- specific’ metrics within a precision rehabilitation approach (Rosenberg et al., 2022).

A further personalization challenge that was identified pertains to the fact that these decisions themselves are individualized, in the sense that personalization is executed in a manner that is distinct for each clinician, grounded in individual knowledge and expertise. However, study participants described how precision approaches could address these concerns by systematizing and standardizing personalization decisions. This finding aligns with the existing literature, which offers examples of integrating data into decision- making processes to enable personalization. For instance, Lelis et al. (2020) have delineated a decision-making framework for generating data-driven recommendations concerning post-cancer rehabilitation interventions, with the objective to prevent and address system-level adverse effects (Lelis et al., 2020). Participants have articulated the necessity of incorporating all pertinent variables into the decision-making algorithms, encompassing not solely performance metrics but also other crucial factors such as motivation and engagement, which have the potential to optimize the intervention’s effectiveness (Cotton et al., 2024). Silverberg and Mikolic present an example in which they discuss the importance of considering individual psychological risk factors and symptoms when making decisions about the clinical management of mild traumatic brain injury. They advocate for integrating psychological principles into the decision-making process (Silverberg & Mikolic, 2023).

Study participants have expressed ethical concerns about making decisions based solely on predictions from large datasets; this concern is rooted in the heterogenous nature of rehabilitation populations, even within specific diagnostic categories. This heterogeneity give rise to inquiries regarding the suitability of AI modeling in precision approaches, underscoring the necessity for elucidating the circumstances under which AI-based predictive models should be employed and delineating the criteria for determining the adequacy of training data. The participants articulated a desire for AI to facilitate the identification of cohorts of individuals with similar characteristics or clinical trajectories. While the participants acknowledged the importance of predictive modeling, their ambivalence suggests the need for precision rehabilitation to delineate the boundary between personalization and standardization. This delineation would establish consensus, procedures, and clear definitions to address concerns regarding over-reliance on algorithmic predictions within rehabilitation’s traditionally individualized approach. Standardized approaches for weighing individual factors in predictive models and clear guidelines for balancing algorithmic predictions with clinical expertise are essential; this will require collaboration between clinicians and data scientists.

In addition, data collection challenges emerged throughout our findings as a barrier to precision rehabilitation. Participants described the significance of incorporating biological, functional, and sociodemographic data into databases that inform precision rehabilitation. The current literature provides examples of how to identify predictive factors using combinations of data sources, including patient-reported outcome measures, functional tests, and biomarkers. For instance, Rushton et al. (2018) delineate this approach to predicting the risk of chronic pain and disability following musculoskeletal injury (Rushton et al., 2018). The type of data used to inform decisions is of interest for precision rehabilitation. Genetic biomarkers are essential for precision medicine. In this study, participants identified the need to better understand the role of genomic biomarkers and to link biological factors to treatment response (Wagner, 2017; Wagner & Kumar, 2019). In their Rehabilomics Research Model, Wagner (2017, 2019) outlines how a genetic focus can intersect with the life course approach to rehabilitation (Wagner, 2017; Wagner & Kumar, 2019).

Regarding the timing and location of data collection, participants expressed concern about current rehabilitation inefficiencies, such as time-consuming assessments that may not capture functionally relevant data or accurately reflect patients’ abilities in their daily lives. Participants describe a key component of the future of precision rehabilitation as optimizing the timing of assessments to coincide with critical decision-making periods in patient trajectories.

Technologies were identified as facilitators of precision assessment and intervention approaches through improved measurement capabilities. Perspectives on facilitators are consistent with those described by Bonato (2021), who identified how wearable sensors and other digital health technologies can improve performance and participation measurement (Bonato, 2021). Participants identified a need to better identify how technologies can help understand and measure patient function. This view parallels French et al.’s response to Cotton’s editorial, where they state: ‘*We do wish to underscore the importance of careful measurement and monitoring of whole-person function as the core of precision rehabilitation*” (French et al., 2022).

However, participants expressed many concerns about the use of technology in precision rehabilitation. About the use of technology for assessment, a significant concern expressed by participants was the use of technology in real-world contexts to reduce the burden of clinic-based data collection. Participants noted that there is still a burden associated with the use of technology because although data is collected without their involvement, they still have to access, monitor, and troubleshoot its use.

In terms of precision nomenclature, participants in this study noted that the term "precision" is not widely used in the rehabilitation field, but that similar concepts are often discussed using different terminology. Thus, there is a clear need to better articulate how precision approaches differ from and align with existing rehabilitation frameworks and terminology. Similarly, French et al. (2022) emphasize the need for deliberate action to realize the potential benefits of precision rehabilitation, including the infrastructure changes necessary to implement the data collection, storage, and analysis requirements of precision rehabilitation (French et al., 2022). This study findings support this assertion, with study participants discussing the need for system-level changes in data storage, multisite data sharing networks, ethical guidelines, data collection using standardized outcome measures, and institutional support for compensated data collection time and for the use of wearable technologies to reduce the burden of clinical assessment. As an example, French et al.’s 2023 describe the potential for leveraging substantial data sets from electronic health records. They articulate a methodology for cultivating a “Precision Rehabilitation Data Repository” for stroke with the goal of incorporating real-world data from wearable devices (French et al., 2023).

### 4.2 Study limitations and next steps for research

The generalizability of the findings is constrained by the inclusion of only three rehabilitation sites, which may be lacking in various institutional cultures, resource contexts, and regional practice variations that could potentially influence perspectives on the implementation of precision rehabilitation. Furthermore, the perspectives of patients/clients were not included into this study. Incorporating these perspectives in future studies will be essential to advancing precision rehabilitation (Cotton et al., 2022; French et al., 2022).

Integration of these qualitative findings with the quantitative scoping study results will be achieved through a convergent mixed methods approach (Pouliot-Laforte et al., 2025) to provide a comprehensive understanding of the current state of precision rehabilitation, gaps in the evidence base, and suggestions for future research.

## 5. Conclusions

This is the first qualitative study of precision rehabilitation to identify key opportunities and challenges. The study’s findings underscore the pivotal emphasis of precision rehabilitation on function and measurement, while concurrently highlighting previously unexplored tensions between standardization and personalization, challenges in the use of technology for data collection, and the need for infrastructure development. The study’s results emphasize function as a cornerstone of precision rehabilitation across measurement (functional biomarkers), interventions (targeted functional outcomes), and systems (optimized rehabilitation trajectories). The analysis of stakeholder perspectives suggests that successful implementation of precision rehabilitation will require balancing data-driven approaches with clinical expertise, developing efficient measurement technologies, and establishing supportive institutional frameworks. These insights can inform the development of implementation strategies and research priorities. They should be considered alongside quantitative evidence evaluating the effectiveness of precision rehabilitation in establishing the next steps in this emerging domain.

## Data Availability

All data produced in the present study are available upon reasonable request to the authors.

## Acknowledgements

The authors wish to thank the study participants as well as the individuals who facilitated study procedures at each of the three participating institutions.

## Contribution list

Conceptualization (APL, DK, DL), Methodology (APL, ED, DL), Formal Analysis (APL, ED, DL), Writing – Original Draft Preparation (APL, DL), Writing – Review and Editing (APL, ED, DK, DL)

## Ethical statement

The study was approved by the Research Ethics Board of CHU Sainte-Justine Research Centre (no. 2024-6324). All study participants provided informed consent prior to their participation.

## References

1. Adams, S. A., & Petersen, C. (2016). Precision medicine : Opportunities, possibilities, and challenges for patients and providers. Journal of the American Medical Informatics Association, 23(4), 787 790. 10.1093/jamia/ocv215

2. Arksey, H., & O’Malley, L. (2005). Scoping studies : Towards a methodological framework. International Journal of Social Research Methodology, 8(1), 19 32. 10.1080/1364557032000119616

3. Bonato, P. (2021). Keynote : Digital Health Technologies and Their Role in the Development of Precision Rehabilitation Interventions (WOS:000684246800041). 200 200. 10.1109/PERCOMWORKSHOPS51409.2021.9431126

4. Braun, V., & Clarke, V. (2006). Using thematic analysis in psychology. Qualitative Research in Psychology, 3(2), 77 101. 10.1191/1478088706qp063oa

5. Buus, N., Nygaard, L., Berring, L. L., Hybholt, L., Kamionka, S. L., Rossen, C. B., Søndergaard, R., & Juel, A. (2022). Arksey and O’Malley’s consultation exercise in scoping reviews : A critical review. Journal of Advanced Nursing, *78*(8), 2304 2312. 10.1111/jan.15265

6. Cotton, R. J., Seamon, B. A., Segal, R. L., Davis, R. D., Sahu, A., McLeod, M. M., Celnik, P., & Ramey, S. L. (2024). *A Causal Framework for Precision Rehabilitation* (arXiv:2411.03919). arXiv. 10.48550/arXiv.2411.03919

7. Cotton, R. J., Segal Rick, R. L., Seamon, B. A., Sahu, A., McLeod, M. M., Davis, R. D., Ramey, S. L., French, M. A., Roemmich, R. T., Daley, K., Beier, M., Penttinen, S., Raghavan, P., Searson, P., Wegener, S., & Celnik, P. (2022). Precision Rehabilitation : Optimizing Function, Adding Value to Health Care. Archives of physical medicine and rehabilitation, 103(9), 1883 1884. 10.1016/j.apmr.2022.04.017

8. Doyle, L., McCabe, C., Keogh, B., Brady, A., & McCann, M. (2020). An overview of the qualitative descriptive design within nursing research. Journal of Research in Nursing: JRN, 25(5), 443 455. 10.1177/1744987119880234

9. Fife, S. T., & Gossner, J. D. (2024). Deductive Qualitative Analysis : Evaluating, Expanding, and Refining Theory. International Journal of Qualitative Methods, 23, 16094069241244856. 10.1177/16094069241244856

10. French, Daley, K., Lavezza, A., Roemmich, R. T., Wegener, S. T., Raghavan, P., & Celnik, P. (2023). A Learning Health System Infrastructure for Precision Rehabilitation After Stroke. American journal of physical medicine & rehabilitation, 102(2S Suppl 1), S56 S60. 10.1097/PHM.0000000000002138

11. French, Roemmich, R. T., Daley, K., Beier, M., Penttinen, S., Raghavan, P., Searson, P., Wegener, S., & Celnik, P. (2022). Precision Rehabilitation : Optimizing Function, Adding Value to Health Care. Archives of Physical Medicine and Rehabilitation, 103(6), 1233 1239. 10.1016/j.apmr.2022.01.154

12. Gambhir, S. S., Ge, T. J., Vermesh, O., & Spitler, R. (2018). Toward achieving precision health. Science Translational Medicine, 10(430), eaao3612. 10.1126/scitranslmed.aao3612

13. Jette, A. (2022). PTJ’s Editor’s Choice. *APTA Magazine*, *14*(2), 48 49.

14. Johnson, K. B., Wei, W.-Q., Weeraratne, D., Frisse, M. E., Misulis, K., Rhee, K., Zhao, J., & Snowdon, J. L. (2021). Precision Medicine, AI, and the Future of Personalized Health Care. Clinical and Translational Science, 14(1), 86 93. 10.1111/cts.12884

15. Lelis, Bolanos J.L., & Wong M. (2020). Matching precision medicine with precision rehabilitation : A framework for clinical decision making in oncology rehabilitation. Rehabilitation Oncology, 38(1), E27. 10.1097/01.REO.0000000000000202

16. Levac, D., Colquhoun, H., & O’Brien, K. K. (2010). Scoping studies : Advancing the methodology. Implementation Science, 5(1), 69. 10.1186/1748-5908-5-69

16. Lin, D. J., & Stein, J. (2023). Stepping Closer to Precision Rehabilitation. JAMA neurology, 80(4), 339 341. 10.1001/jamaneurol.2023.0044

17. Patton, M. (2014). Qualitative Research & Evaluation Methods : Integrating Theory and Practice (4th edition). Sage Publications.

18. Pouliot-Laforte, A., Dubé, E., Kairy, D., & Levac, D. E. (2025). What is the current state of precision rehabilitation? Protocol for a scoping study with a consultation phase. BMJ Open, 15(1), e094119. 10.1136/bmjopen-2024-094119

19. Rosenberg, Christianson H., Liu J., Santucci V., Sims P., Schilder A., Zajac-Cox L., Winner T.S., Ting L.H., & Kesar T.M. (2022). Fastest may not be the best : Differential and individual-specific immediate effects of gait speed on biomechanical variables post- stroke. *medRxiv*, *(Rosenberg, Winner, Ting) Department of Biomedical Engineering, Emory University, Georgia Institute of Technology, Atlanta, GA, United States(Christianson, Liu, Santucci, Sims, Schilder, Zajac-Cox, Kesar) Department of Rehabilitation Medicine, Emory Unive*. 10.1101/2022.12.14.22283438

20. Rushton, A. B., Evans, D. W., Middlebrook, N., Heneghan, N. R., Small, C., Lord, J., Patel, J. M., & Falla, D. (2018). Development of a screening tool to predict the risk of chronic pain and disability following musculoskeletal trauma : Protocol for a prospective observational study in the United Kingdom. BMJ open, *8*(4), e017876. 10.1136/bmjopen-2017-017876

21. Sandelowski, M. (2010). What’s in a name? Qualitative description revisited. Research in Nursing & Health, 33(1), 77 84. 10.1002/nur.20362

22. Schleidgen, S., Klingler, C., Bertram, T., Rogowski, W. H., & Marckmann, G. (2013). What is personalized medicine : Sharpening a vague term based on a systematic literature review. BMC Medical Ethics, 14(1), 55. 10.1186/1472-6939-14-55

23. Silverberg, N. D., & Mikolic, A. (2023). Management of Psychological Complications Following Mild Traumatic Brain Injury. Current neurology and neuroscience reports, 23(3), 49 58. 10.1007/s11910-023-01251-9

24. Tong, A., Sainsbury, P., & Craig, J. (2007). Consolidated criteria for reporting qualitative research (COREQ) : A 32-item checklist for interviews and focus groups. International Journal for Quality in Health Care, 19(6), 349 357. 10.1093/intqhc/mzm042

25. Vaismoradi, M., Turunen, H., & Bondas, T. (2013). Content analysis and thematic analysis : Implications for conducting a qualitative descriptive study. Nursing & Health Sciences, 15(3), 398 405. 10.1111/nhs.12048

26. Wagner, A. K. (2017). TBI Rehabilomics Research : An Exemplar of a Biomarker- Based Approach to Precision Care for Populations with Disability. Current neurology and neuroscience reports, 17(11), 84. 10.1007/s11910-017-0791-5

26. Wagner, A. K., & Kumar, R. G. (2019). TBI Rehabilomics Research : Conceptualizing a humoral triad for designing effective rehabilitation interventions. Neuropharmacology, *145*(Pt B), 133 144. 10.1016/j.neuropharm.2018.09.011

27. World Health Organization. (2023). *Rehabilitation*. Rehabilitationf. https://www.who.int/news-room/fact-sheets/detail/rehabilitation

